# Racial Differences in the Relationship Between Blood Pressure and Cognitive Decline

**DOI:** 10.1101/2024.01.03.24300811

**Authors:** Michael D. Oliver, Cassandra Morrison, Sondos El-Hulu, Marquinta Harvey, Lisa L. Barnes

**Author notes:** Corresponding author: Michael D. Oliver, Department of Psychological Science and Neuroscience, Belmont University, 1900 Belmont Blvd. Nashville, TN, 37212.

## Abstract

**Background:** Cognition may be influenced by health-related factors such as blood pressure (BP). However, variations in BP may differentially affect cognition as a function of race. This study investigates the relationship between normal, high, and variable BP and cognitive decline in older Black and White adults.

**Methods:** 2284 participants (1139 Blacks, 1145 Whites, *M*_Age_=73.4, *SD*=6.6) from 3 harmonized cohorts of older adults from the Rush Alzheimer’s Disease Center, were divided into 3 groups (normal, high, variable) based on systolic BP mean and standard deviation. Cognitive scores were computed from multiple assessments in 5 domains (i.e., episodic memory, semantic memory, working memory, processing speed, visuospatial ability). Performance across 19 tests were averaged to create a measure of global cognition. Linear mixed-effects models examined racial differences between BP and cognitive change over an average of 6.7 years.

**Results:** White adults with high or variable BP had faster rates of decline in global cognition compared to Black adults. White adults with high BP declined faster in perceptual speed, semantic memory, and working memory compared to Black adults with high BP, whereas White adults with variable BP had faster rates of decline in all cognitive domains compared to Black adults with variable BP. No racial differences were observed in individuals with normal BP.

**Conclusions:** White older adults with elevated or fluctuating BP show faster rates of cognitive decline compared to older Black adults. Findings highlight the complex interplay between BP and cognitive health, emphasizing the need for targeted interventions to address racial disparities in cognitive well-being.

## Introduction

According to the World Health Organization, hypertension is estimated to affect nearly 1.3 billion people worldwide^1^. Several studies have revealed inverse associations between blood pressure (BP) and cognition^2–4^ and have identified hypertension as a key modifiable risk factor for cognitive decline secondary to Alzheimer’s disease (AD) and related dementia.^5^ However, the extent to which hypertension affects cognitive abilities may differ as a function of race. It has been well-documented that the prevalence of hypertension is higher in minorities compared to Whites.^6, 7^ This increased prevalence contributes to higher cumulative BP levels that may explain disparities in cognitive abilities. For example, a pooled cohort analysis of 19,378 participants from 5 different studies, revealed that Black individuals with higher cumulative BP levels demonstrated significantly faster decline in memory and global cognition compared to White individuals.^8^ Moreover, not only are cumulative BP rates typically higher, but control rates tend to be significantly lower in Black individuals compared to White individuals.^7, 9^ These differences in BP values and control undoubtedly contribute to differences in the way that BP influences cognitive abilities over time. Although Black individuals report higher cumulative BP, and higher BP is thought to be suggestive of greater cognitive decline, the effects that BP has on specific cognitive abilities stratified by race may not be as clear. As such, the relationship between BP and cognitive abilities as a function of race warrants further investigation. Identifying, targeting, and treating those with high BP proves to be a critical step in minimizing risk of cognitive decline.

Few studies have explored BP-related changes in cognitive abilities using long follow-up times and multiple cognitive domains. Furthermore, few studies have examined the influence of variable BP on cognition, and if this relationship differs by race. Therefore, the current study aims to investigate race differences in the relationship between BP and longitudinal cognitive performance. This work may allow for targeted therapeutic interventions for subsequent care planning specific to racial/ethnic group.

## Method

### Participants

Data were obtained from three harmonized cohort studies on aging and dementia: Minority Aging Research Study (MARS)^10^, Rush Memory and Aging Project (MAP)^11^, and Black Clinical Core^12^. These studies have essentially identical recruitment techniques, testing procedures, and a large overlap of data collection at the item level by RAs trained by the same trainer. This method allows for data to be merged seamlessly for analyses. Annual clinical evaluations were conducted face-to-face in participants’ homes and included a medical history, neurological examination, BP measurement and cognitive testing (described below). Written informed consent was obtained from all participants, and research was approved by an Institutional Review Board of Rush University Medical Center. Secondary analyses included in this paper were approved by the Belmont University Institutional Review Board. More information about each cohort study design can be accessed online through the RADC Research Resource Sharing Hub (https://www.radc.rush.edu/docs/parentStudyDesigns.htm).

Participant inclusion criteria for this study were as follows: 1) at least 55 years of age at baseline, 2) self-report as Black or White, 3) had completed at least two cognitive assessments, and 4) had at least two BP readings completed at their annual visits. A total of 2284 participants met the above-mentioned inclusion criteria were included in this study.

### Blood Pressure Readings

Mean systolic blood pressure (SBP) and standard deviation (SD) was computed for each individual for all timepoints. That is, all of each participant’s blood pressure readings across all timepoints were averaged to create a mean SBP for each person. The SD was then computed for the entire sample. Participants were then divided into the following 3 groups based on mean and SD: 1) *normal BP* – mean BP had to be below 130 and SD not more than 1 SD away from the sample SD, 2) *high* BP– mean BP had to be greater than or equal to 130 (as per the National Institute of Health and National Institute on Aging guidelines for older adults) and SD not more than 1 SD away from sample SD^13^, and 3) *variable BP*– BP SD was more than 1 SD away from the sample SD. The 2284 participants were divided into one of three blood pressure groups, normal, high, or variable. There was therefore a total of 6 groups, White participants – normal BP (n = 396), White participants – high BP (n = 332), White participants – variable BP (n = 416), Black participants – normal BP (n = 259), Black participants – high BP (n = 351), Black participants – variable BP (n = 529).

### Cognitive Assessment

All participants were administered a battery of neuropsychological tests including 19 tests selected to assess five cognitive domains.^14, 15^ There were seven tests of episodic memory (immediate and delayed recall of Story A of the Wechsler Memory Scale-Revised; immediate and delayed recall of the East Boston Story; Word List Memory, Recall and Recognition), three tests of semantic memory (Verbal Fluency; Boston Naming; Reading Test), three tests of working memory (Digit Span forward and backward; Digit Ordering), four tests of processing speed (Symbol Digit Modalities Test; Number Comparison; two indices from a modified version of the Stroop Test), and two tests of visuospatial ability (Line Orientation; Progressive Matrices). Composite measures of each domain were used in analyses. To create each composite score, individual tests were converted to z-scores, using the mean and standard deviation from the combined cohort at baseline, and z-scores for the relevant tests were averaged. An individual’s standard performance across all 19 of these tests was averaged to create a measure of global cognitive function.^16^ More information for the specific tests used for each category can be obtained from https://www.radc.rush.edu/.

### Statistical Analysis

Analyses were performed using “R” software version 4.0.5. Participants were grouped into one of six groups based on their race (either Black or White) and BP status (normal, high, variable).

Linear mixed effects models (“lmer”, package “lme4” in R) were used to examine cognitive change over time. Analyses were completed for each cognitive domain of interest (global functioning, episodic memory, visuospatial abilities, processing speed, semantic memory, and working memory), and the interaction between Time-From-Baseline and Group was examined. The first analysis was completed with White normal BP as the reference, because there were two other White groups to compare the Black group to, the analyses were repeated twice using White high BP and White variable BP as the reference. Participant ID was included as a categorical random effect to account for repeated measures of the same participant.

The model also included age at baseline, sex, years of education, and baseline diagnosis (categorical variable contrasting MCI, dementia, and AD against the controls) as covariates.

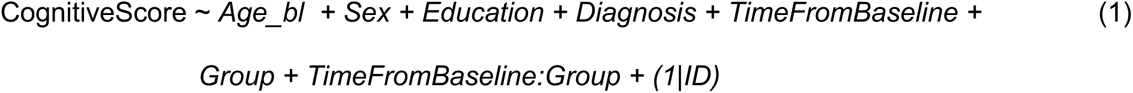

All models were corrected for multiple comparisons using false discovery rate (FDR)^17^, due to presence of interdependency between the regression and mixed effects models performed. All p-values were reported as raw values with significance, then determined by FDR correction. All continuous values were z-scored within the population prior to the analyses. For the purposes of this paper, we were interested in comparing each White group to the three Black groups. Thus, within race-group comparisons are not reported.

To ensure that any race differences observed were not due to differences in age, sex, education, or diagnostic differences between the groups, the same number of White individuals were selected based on matching criteria to the Black sample. The groups were matched based on sex, diagnosis, age, and years of education. Participant demographic information by group is presented in Table 1.

**Table 1:** Demographic table by race and by group.

|  | Black | White | Black Normal<br>BP | Black High BP | Black Variable<br>BP | White Normal<br>BP | White High BP | White Variable<br>BP |
| --- | --- | --- | --- | --- | --- | --- | --- | --- |
| Sample (females) | 1139 (79%) | 1145 (78%) | 259 (82%) | 351(75%) | 529 (81%) | 396 (83%) | 332 (77%) | 416 (74%) |
| Baseline Diagnosis |  |  |  |  |  |  |  |  |
| NC | 845 (75%) | 868 (76%) | 194 (75%) | 268 (76%) | 383 (72%) | 306 (77%) | 250 (75%) | 311 (75%) |
| MCI | 258 (22%) | 238 (21%) | 57 (22%) | 73 (21%) | 128 (24%) | 76 (20%) | 74 (22%) | 87 (21%) |
| AD | 32 (3%) | 35 (3%) | 8 (3%) | 9 (3%) | 15 (3%) | 12 (3%) | 5 (2%) | 18 (4%) |
| Dementia | 4 (<1%) | 5 (<1%) | 0 (0%) | 1 (<1%) | 3 (<1%) | 2 (<1%) | 3 (1%) | 0 (0%) |
| Age | 73.2 ± 6.6 | 73.6 ± 6.6 | 72.4 ± 6.8 | 72.7 ± 6.6 | 74.0 ± 6.3 | 72.4 ± 6.7 | 73.8 ± 6.5 | 74.5 ± 6.5 |
| Education | 15.0 ± 3.5 | 15.4 ± 3.8 | 15.0 ± 3.4 | 15.0 ± 3.4 | 15.0 ± 3.5 | 15.7 ± 3.9 | 15.5 ± 3.7 | 14.9 ± 3.8 |
| Mean BP | 136.1 ± 15.3 | 130.6 ± 14.1 | 119.81 ± 6.7 | 143.1 ± 12.0 | 139.5 ± 14.6 | 118.4 ± 7.7 | 139.9 ± 10.5 | 135.1 ± 13.1 |
| Global cognition | -0.20 ± 0.76 | 0.17 ± 0.83 | -0.16 ± 0.84 | -0.16 ± 0.75 | -0.24 ± 0.72 | 0.23 ± 0.85 | 0.20 ± 0.78 | 0.09 ± 0.85 |
| Episodic memory | -0.18 ± 0.75 | -0.01 ± 0.84 | -0.18 ± 0.81 | -0.15 ± 0.76 | -0.20 ± 0.71 | 0.06 ± 0.82 | -0.01 ± 0.78 | -0.08 ± 0.90 |
| Visuospatial ability | -0.35 ± 0.97 | 0.24 ± 0.89 | -0.31 ± 1.04 | -0.27 ± 0.95 | -0.42 ± 0.95 | 0.30 ± 0.98 | 0.28 ± 0.81 | 0.16 ± 0.88 |
| Processing speed | -0.08 ± 0.75 | 0.30 ± 0.91 | 0.01 ± 0.81 | -0.07 ± 0.76 | -0.14 ± 0.72 | 0.35 ± 0.96 | 0.34 ± 0.89 | 0.22 ± 0.90 |
| Semantic memory | -0.15 ± 0.86 | 0.23 ± 0.81 | -0.12 ± 0.94 | -0.10 ± 0.88 | -0.20 ± 0.81 | 0.27 ± 0.84 | 0.22 ± 0.75 | 0.19 ± 0.82 |
| Working memory | -0.14 ± 0.99 | 0.17 ± 0.99 | -0.17 ± 0.93 | -0.11 ± 0.90 | -0.15 ± 0.86 | 0.16 ± 0.99 | 0.26 ± 1.01 | 0.10 ± 0.98 |

Sample = total sample size (percentage of females). Mean BP = mean BP at all measurement time points. BP= blood pressure. BL = baseline. Cognitive scores represent the averaged baseline z-scores by group.

## Results

### Cognitive Change

Figure 1 shows the mixed effects model predictions of cognitive scores over time for each cognitive domain by group. As expected, for global function, and all cognitive domains (episodic memory, visuospatial abilities, processing speed, semantic memory, and working memory), Time From Baseline (*t* belongs to [-6.26 to -18.28], *p*<.001) and increased age at baseline (*t* belongs to [-5.09 to -18.61], *p*<.001) were associated with increased rate of cognitive change. All results remained statistically significant after FDR correction.

**Figure 1:**
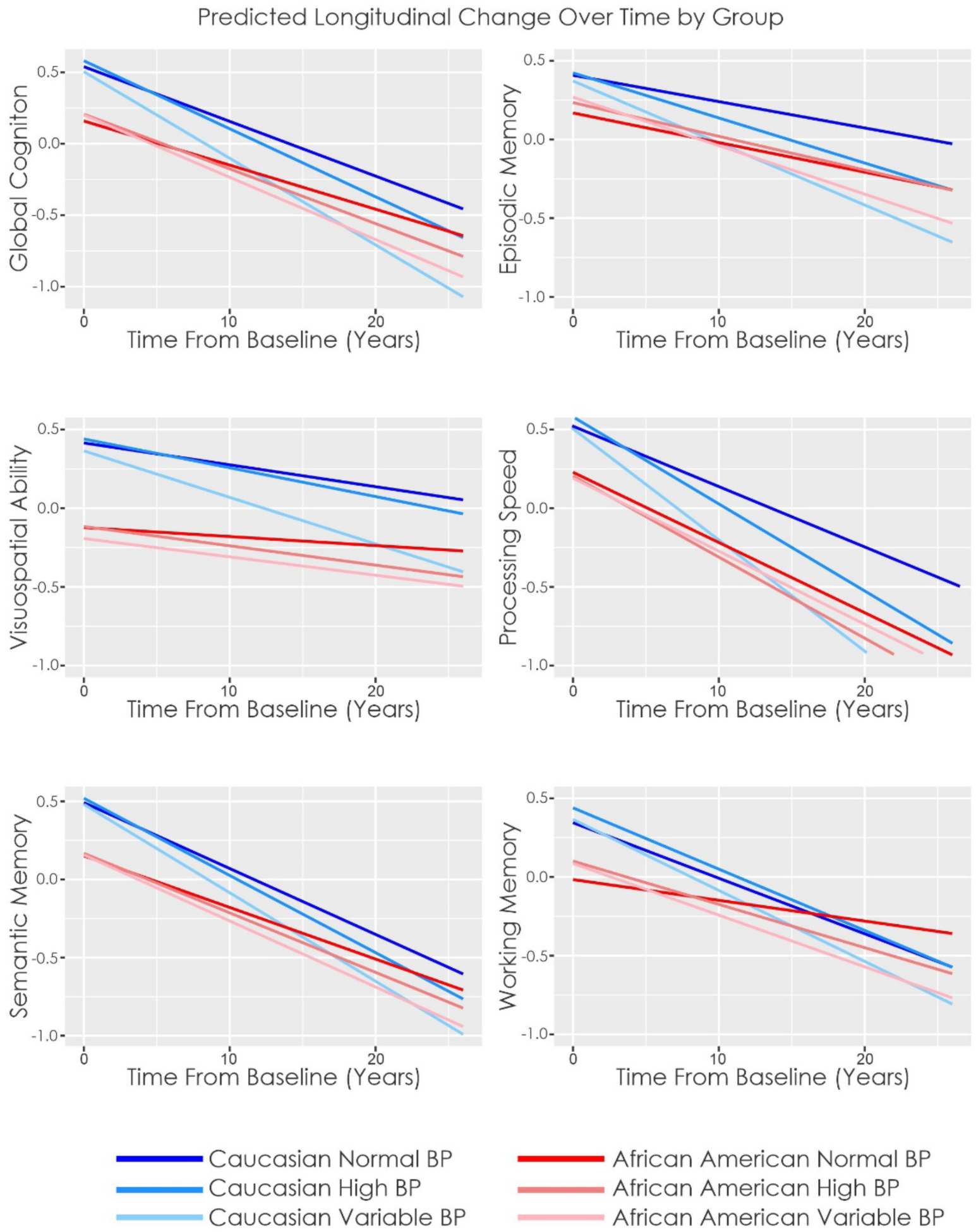
Predicted Longitudinal Change Over Time by Group. Longitudinal change over time in White and Blacks with normal, high, and variable blood pressure. Plotted cognitive scores are z-scored values. BP = blood pressure.

For global cognition, the interaction between Time From Baseline and Group showed that White normal BP rate of cognitive change did not differ from any of the three Black groups (normal, high, and variable BP). The White high BP group had increased rate of decline compared to Black normal and high BP (*t* belongs to [-2.13 to -3.98], *p*<.05), but not Black variable BP. The White variable BP group had increased rate of decline compared to all three Black groups (normal, high, and variable BP), (*t* belongs to [-5.49 to -7.67], *p*<.001).

For episodic memory, the White normal BP groups rate of cognitive change was less than that of Black Variable BP (*t* = 3.86, *p*<.001), but did not differ from Black normal or high BP. After correction for multiple comparisons, White high BP did not differ from any of the Black groups. The White variable BP group had increased rate of decline compared to all three Black groups (normal, high, and variable BP), (*t* belongs to [-2.48 to -4.69], *p*<.05).

For visuospatial abilities, White normal BP rate of cognitive change did not differ from any of the three Black groups (normal, high, and variable BP). The White high BP group had an increased rate of decline compared to Black normal BP (*t* = -2.39, *p* = .017). The White variable BP group had increased rate of decline compared to all three Black groups (normal, high, and variable BP), (*t* belongs to [-3.39 to -4.89], *p*<.001).

For processing speed, White normal BP rate of cognitive change was less than that of only Black high BP (*t =* 2.26, *p*=.024). The White high BP group had increased rate of decline compared to Black normal and high BP (*t* belongs to [-1.03 to -2.76], *p*<.05). The White variable BP group had increased rate of decline compared to all three Black groups (normal, high, and variable BP), (*t* belongs to [-4.80 to -8.11], *p*<.001).

For semantic memory, White normal BP rate of cognitive change did not differ from any of the three Black groups (normal, high, and variable BP), after correction for multiple comparisons. The White high BP group exhibited an increased rate of decline compared to Black normal and high BP (*t* belongs to [-2.28 to -3.43], *p*<.05). The White variable BP group had increased rate of decline compared to all three Black groups (normal, high, and variable BP), (*t* belongs to [-4.01 to -5.35], *p*<.001).

For working memory, White normal BP rate of cognitive change was more than that of only Black normal BP (*t* = -4.90, *p*<.001). The White high BP group had increased rate of decline compared to Black normal and high BP (*t* belongs to [-2.32 to -5.44], *p*<.05). The White variable BP group had increased rate of decline compared to all three Black groups (normal, high, and variable BP), (*t* belongs to [-3.56 to -7.31], *p*<.001).

It should be noted that the RUSH dataset contains data from multiple cohorts. To ensure results were not driven by cohort effects, we repeated all analysis with cohort as a random categorical effect (1| Cohort). For a more conservative assessment, we also repeated the models including Time From Baseline as a random slope: (1 + TimeFromBaseline| ID). In both analyses, all results remained the same in terms of effect size and significance.

### Secondary Exploratory Analyses

One possible explanation for Black older adults having lower rates of decline is associated with lower cognitive scores at baseline compared to White older adults. To test this possibility, we reran all analyses comparing “high” and “low” performers collapsed across race and separated each cognitive domain by a median split. These two groups were then divided into normal, high, and variable BP following our previous methods. Using the same analyses (equation 1), we observed that the low performers with all BP types had steeper declines in cognition over time compared to high performers of all BP types (see supplementary material Table 1) in all domains except visuospatial ability and working memory. In Visuospatial ability, none of the low performer groups differed from high performers in rate of cognitive change. In working memory, the low performers with high and variable BP types had steeper declines in cognition compared to high performers of all BP types; however, low performers with normal BP did not differ from high performers of any BP type in rate of cognitive change. Therefore, the reduced association between BP and cognitive change in older Black adults is not associated with lower baseline cognitive scores.

## Discussion

According to a systematic review of 50 studies including 107,405 participants, high BP plays a significant role in cognitive impairment.^18^ Controlling BP using medications and other treatments has been shown to be successful in mitigating the negative effects of BP on cognition^19^; however, what is not well-understood is whether fluctuations in BP are associated with long-term cognitive changes. Moreover, although the relationship between BP and cognition is well-documented in the literature, whether this relationship differs as a function of race is less understood. As such, this study investigated the effects of BP on cognitive decline by race. Specifically, we examined relationships between normal, high, and variable BP on domain-specific and global cognitive decline separately in older Black and White adults. Findings from this study reveal no racial differences in the rate of cognitive decline among individuals with normal BP. White adults with high or variable BP exhibited significantly faster rates of decline in global cognition compared to their Black counterparts. More specifically, White adults with high BP declined faster in perceptual speed, semantic memory, and working memory compared to Black adults with high BP, whereas White adults with variable BP had significantly faster rates of decline in all five cognitive domains when compared to Black adults with variable BP.

Findings from this study show that in individuals with normal BP, the influence of BP on rate of cognitive decline does not differ between Black and White older adults. That is, when comparing Blacks and Whites with similar mean BP within the normal range, BP does not differentially impact one racial group more than the other. Our findings are consistent with another study that found no differences in cognitive decline when comparing cognitive aging in Black and White older adults over 5 years.^20^ Moreover, one study observed that race differences in cognitive performance were no longer significant when BP was controlled for in the analyses.^8^ As such, given that Blacks have a higher prevalence of hypertension, undoubtedly, risk of cognitive decline may be higher as a direct result. However, this increased risk of cognitive decline may not be specific to Black people, because in people with normal BP, the relationship between BP and cognition does not seem to vary as a function of race. Taken together, maintaining and promoting healthy BP across race may result in the elimination of any race-related differences in how BP impacts cognitive abilities over time.

Elevated BP is a significant risk factor for several health conditions, including those affecting cognitive abilities such as Alzheimer’s disease and related dementias.^21^ The present study’s findings revealed that in individuals with high BP, older Black and White adults had similar rates of decline in episodic memory and visuospatial abilities. However, race differences were present in the rate of decline in those with high BP in global cognition, perceptual speed, semantic memory, and working memory; all of which White individuals exhibited significantly faster rates of cognitive decline than Black individuals. Our findings are consistent with a recent MRI study linking high BP to brain changes in specific structural regions (i.e., the putamen, anterior thalamic radiation, anterior corona radiata, and anterior limb of the internal capsule) to impaired executive function, processing speed, learning.^22^ However, in our study, we find that these functional abilities localized to primarily frontal cortical areas, affect White individuals more than Black individuals. Therefore, although we see similar effects of BP on cognitive abilities to those observed previously^22^, the deleterious effects of high BP are found to differentially impact cognitive performance in White compared to older Black adults. Moreover, Whites with variable BP had significantly faster rates of decline in global cognitive abilities, as well as all domain-specific cognitive performance (i.e., episodic memory, visuospatial abilities, perceptual speed, semantic memory, working memory), when compared to Blacks with variable BP.

Another reason for these disparities in BP’s effect on cognitive abilities may be explained by the functional status of the autonomic nervous system (ANS) – the branch of the peripheral nervous system that regulates BP. For example, the ANS regulates BP via vasoconstriction as BP rises, and vasodilation as BP drops. More specifically, the sympathetic branch of the ANS is dominant as BP is elevated causing blood vessels to vasoconstrict which increases BP. Prolonged sympathetic nervous system (SNS) activation can lead to sustained high BP^23^, which may ultimately negatively affect cognitive function.^24^ Fluctuations in BP may be associated with an impaired functioning ANS, or homeostatic imbalance. Homeostatic imbalance reflects the inability to regulate SNS activity, and it has been linked to several conditions considered risk factors for AD (i.e., cardiovascular disease, hypertension, diabetes).^25^ For example, research shows that parasympathetic dysfunction is evident in the prodromal stages of the disease (i.e., mild cognitive impairment).^26^ Moreover, autonomic dysregulation has also been observed in 66% of individuals with AD.^27^ Therefore, ANS functional changes may represent changes in cognitive abilities reflective of AD pathology that can be measured in normal body functioning such as BP.

It Is thought that prolonged SNS activation may be indicative of sustained increases in allostatic load. Allostatic load refers to the cumulative “wear and tear” of chronic stress and life events.^27^ Higher allostatic loads and chronic stress are associated with key indicators of accelerated aging, psychological well-being, and slow deterioration of cognitive functioning.^29.30^ However, it is possible that continuous prolonged activation of the SNS due to various life course social factors may result in a diminished or weakened response to increases in allostatic load. Black individuals experience a disproportionate burden of racism-related stress and discrimination associated with poverty, employment, and residential segregation compared to White individuals.^31, 32^ This chronic stress may cause dysregulation in SNS, hypothalamic-pituitary-adrenal (HPA) axis, immune system, and in cardiovascular and metabolic processes.^33^ As a result, excess SNS activity may act to weaken the body’s immune system, thus leading to a higher prevalence of disease such as cardiovascular disease, hypertension, stroke, heart attacks, and diabetes; all of which are more prevalent in Black individuals.^34^ Over time, frequent exposures to factors increasing allostatic load may have a lesser impact on cognitive function in Black individuals, potentially due to strengthened neural connections. The automatic processing of these experiences, without rumination, may contribute to a weakening effect on cognitive function. Greater lifetime exposures to allostatic load factors, such as racism-related stress and discrimination, may result in older Black adults being less affected cognitively by stress-induced physiological changes like elevated blood pressure compared to older White adults. Our findings support that older Black adults may experience less cognitive impact from stress-induced physiological changes, such as elevated blood pressure, in comparison to their White counterparts.

Although the current study is novel in that it identifies race differences in the relationship between BP and cognition, there are a couple limitations to note. First, the present study operationalized BP into three distinct categories based on individual yearly means and standard deviations compared to the entire sample. Although this technique provides an objective measure of BP that can be compared across the sample, it does not consider factors such as adherence to medication, which may influence BP. For example, it has been well-documented that adherence to pharmacologic treatment of BP can lead to more controlled and/or reduced BP levels compared to low or no adherence.^35, 36^ Therefore, although this does not change the results of the present study, evaluating and tracking medication adherence may influence specific BP categories that individuals may be categorized in. Future studies may wish to assess medication adherence as a potential covariate in the relationship between BP and cognition. Second, a comprehensive history of medication use was not included in the present study. Previous research shows that various classes of medications may impair cognitive abilities in older adults.^37–40^ Although beyond the scope of this study, a detailed exploration into medication use would be beneficial as it may provide additional insights into the various medications that may influence cognitive abilities. Although these limitations are present, overall, the current study was novel in that it examined the relationship between normal, high, and variable BP on cognition in older White and Black adults. These findings reveal a significant disparity, highlighting that older White adults exhibit a higher susceptibility to cognitive decline when facing elevated and fluctuating BP levels, as compared to their Black counterparts. These results shed light on the complex interplay between BP and cognitive health, emphasizing the need for targeted interventions to address racial disparities in cognitive well-being.

## Data Availability

All data produced are available online at the RADC Research Resource Sharing Hub. To obtain data from MARS, AA Core, and MAP for research use, please visit the RADC Research Resource Sharing Hub (www.radc.rush.edu).

## Acknowledgements

We want to acknowledge all the MARS, AA Core, and MAP participants. We are also grateful for the hard work from the staff and investigators at the Rush Alzheimer’s Disease Center. To obtain data from MARS, AA Core, and MAP for research use, please visit the RADC Research Resource Sharing Hub (www.radc.rush.edu).

---

**Supplemental Table 1:**
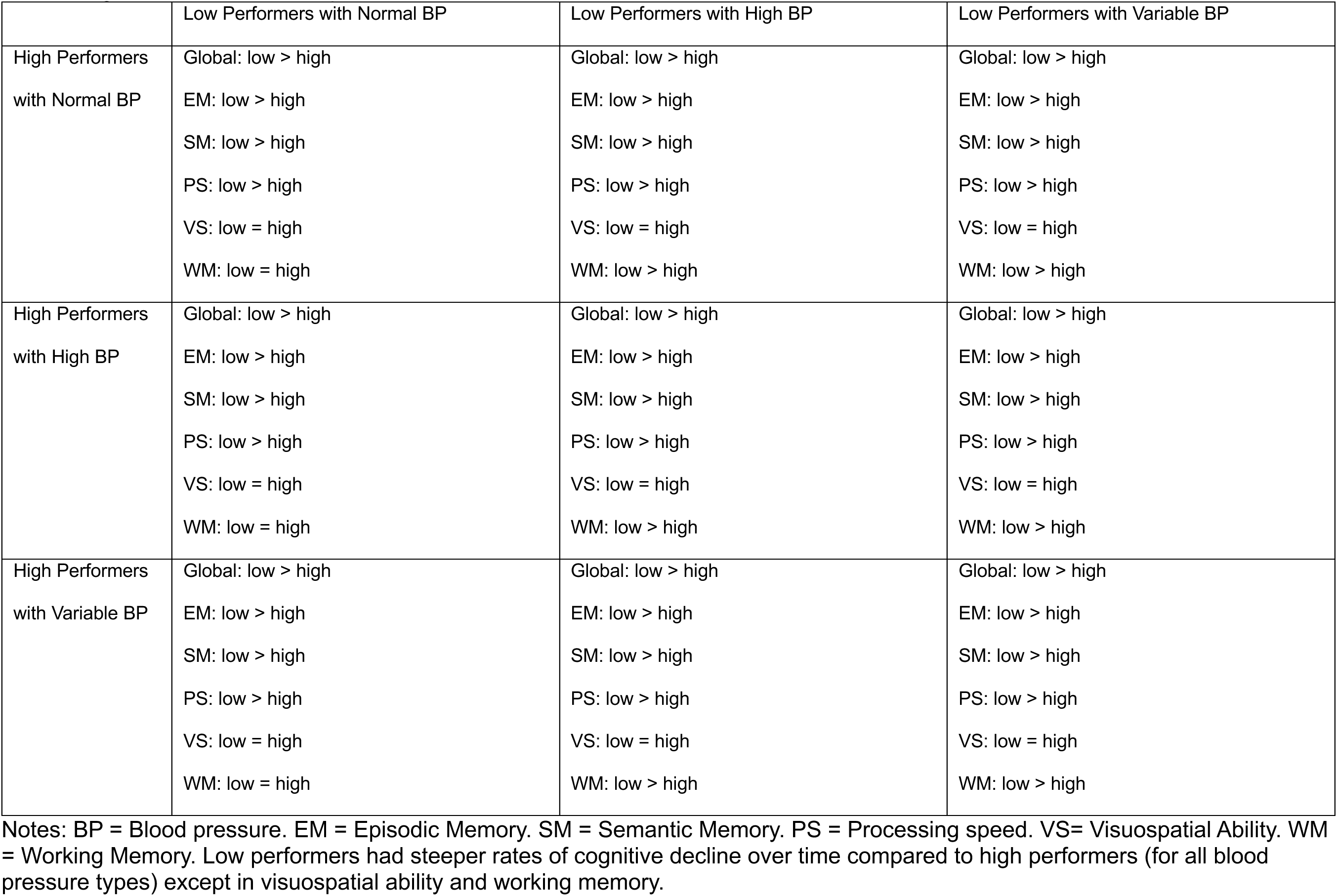
Showing the linear mixed effects results in cognitive change over time between high and low performers with normal, high, and variable blood pressure.

|  | Low Performers with Normal BP | Low Performers with High BP | Low Performers with Variable BP |
| --- | --- | --- | --- |
| High Performers<br>with Normal BP | Global: low > high<br>EM: low > high<br>SM: low > high<br>PS: low > high<br>VS: low = high<br>WM: low = high | Global: low > high<br>EM: low > high<br>SM: low > high<br>PS: low > high<br>VS: low = high<br>WM: low > high | Global: low > high<br>EM: low > high<br>SM: low > high<br>PS: low > high<br>VS: low = high<br>WM: low > high |
| High Performers<br>with High BP | Global: low > high<br>EM: low > high<br>SM: low > high<br>PS: low > high<br>VS: low = high<br>WM: low = high | Global: low > high<br>EM: low > high<br>SM: low > high<br>PS: low > high<br>VS: low = high<br>WM: low > high | Global: low > high<br>EM: low > high<br>SM: low > high<br>PS: low > high<br>VS: low = high<br>WM: low > high |
| High Performers<br>with Variable BP | Global: low > high<br>EM: low > high<br>SM: low > high<br>PS: low > high<br>VS: low = high<br>WM: low = high | Global: low > high<br>EM: low > high<br>SM: low > high<br>PS: low > high<br>VS: low = high<br>WM: low > high | Global: low > high<br>EM: low > high<br>SM: low > high<br>PS: low > high<br>VS: low = high<br>WM: low > high |

Notes: BP = Blood pressure. EM = Episodic Memory. SM = Semantic Memory. PS = Processing speed. VS= Visuospatial Ability. WM = Working Memory. Low performers had steeper rates of cognitive decline over time compared to high performers (for all blood pressure types) except in visuospatial ability and working memory.

**Supplemental Figure 1:**
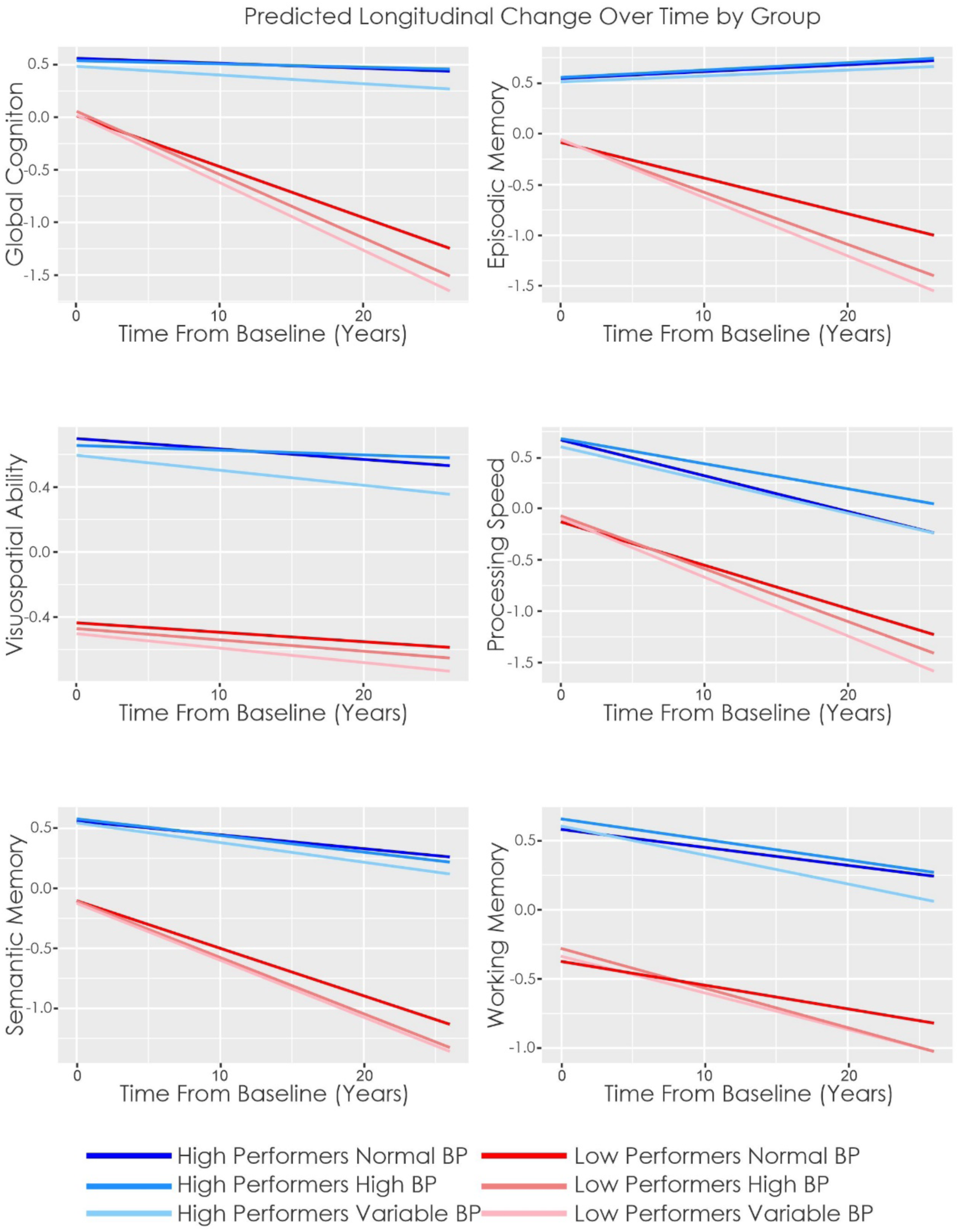
Predicted Longitudinal Change Over Time by Group.

